# Probing Large Language Model Hidden States for Adverse Drug Reaction Knowledge

**DOI:** 10.1101/2025.02.09.25321620

**Authors:** Jacob Berkowitz, Davy Weissenbacher, Apoorva Srinivasan, Nadine A. Friedrich, Jose Miguel Acitores Cortina, Sophia Kivelson, Graciela Gonzalez Hernandez, Nicholas P. Tatonetti

**Affiliations:** Department of Computational Biomedicine, Cedars-Sinai Medical Center, Los Angeles, California, USA; Cedars-Sinai Cancer, Cedars-Sinai Medical Center, Los Angeles, California, USA

**Keywords:** Interpretable AI, Sparse Autoencoder, Knowledge Synthesis, Adverse drug reactions

## Abstract

Large language models (LLMs) integrate knowledge from diverse sources into a single set of internal weights. However, these representations are difficult to interpret, complicating our understanding of the models’ learning capabilities. Sparse autoencoders (SAEs) linearize LLM embeddings, creating monosemantic features that both provide insight into the model’s comprehension and simplify downstream machine learning tasks. These features are especially important in biomedical applications where explainability is critical. Here, we evaluate the use of Gemma Scope SAEs to identify how LLMs store known facts involving adverse drug reactions (ADRs). We transform hidden-state embeddings of drug names from Gemma2-9b-it into interpretable features and train a linear classifier on these features to classify ADR likelihood, evaluating against an established benchmark. These embeddings provide strong predictive performance, giving AUC-ROC of 0.957 for identifying acute kidney injury, 0.902 for acute liver injury, 0.954 for acute myocardial infarction, and 0.963 for gastrointestinal bleeds. Notably, there are no significant differences (p > 0.05) in performance between the simple linear classifiers built on SAE outputs and neural networks trained on the raw embeddings, suggesting that the information lost in reconstruction is minima. This finding suggests that SAE-derived representations retain the essential information from the LLM while reducing model complexity, paving the way for more transparent, compute-efficient strategies. We believe that this approach can help synthesize the biomedical knowledge our models learn in training and be used for downstream applications, such as expanding reference sets for pharmacovigilance.

## 1 Introduction

Through training, large language models (LLMs) synthesize information from diverse sources into coherent representations. Large pretrained models like OpenAI’s GPT-4[1], Google’s Gemma[2], etc. have demonstrated exceptional abilities in understanding context and generating human-like text, making them valuable for advancing scientific research across various domains [3].

Evaluating large language models’ (LLMs) knowledge, or probing [4], solely through generation tasks can be misleading, as these tasks may not reveal the model’s true internal knowledge representation. The GPT-4 technical report [1] highlights that current evaluation methods can lead to models appearing overconfident while producing inaccurate predictions [5]. This systematic bias suggests we need more sophisticated approaches to probe and understand LLM capabilities.

One promising approach is to classify based on residual stream activations in the hidden layers of LLMs. This approach considers the internal “*hidden state*” layers of the model, which may retain more accurate signals of confidence and correctness than the output layer [6]. However, a notable challenge in this approach is the issue of superposition, where features are not linearly separable and often polysemantic, responding to mixtures of unrelated inputs [7, 8]. While deep learning approaches can address this complexity, they often lack explainability and are computationally intensive[9].

Sparse autoencoders (SAEs) offer a compelling solution for extracting monosemantic features, or features dedicated to a single and specific concept, from LLM residual streams by transforming complex activations into interpretable components. The process involves transforming the activations to include a sparsity constraint on the internal activations. This encourages most neurons to remain inactive while a select few, termed feature neurons, become highly active. These active neurons are designed to represent isolated concepts, promoting monosemanticity and providing a clearer window into the model’s understanding[8, 10].

We propose using this approach to evaluate LLM knowledge of adverse drug reactions (ADRs) for given drugs, an important task in pharmacovigilance. By using SAEs to distill LLM embeddings into interpretable features, we can improve our understanding of the biomedical knowledge embedded in these models.

ADRs are unfavorable reactions associated with drug use, whether preventable or not [11], and represent a significant concern in patient care, often contributing to increased morbidity, hospitalizations, and healthcare costs[12–14]. Despite advancements in pharmacovigilance, identifying and characterizing ADRs remains challenging due to the fragmented and unstructured nature of relevant data, including clinical trials, electronic health records, and social media platforms[15–17].

Current methods for ADR detection rely heavily on natural language processing (NLP) tools[18, 19], which may fail to capture the complex relationships between drugs and adverse effects. Traditional pharmacovigilance systems also depend on spontaneous reporting, which is frequently hurt by underreporting, delays, and incomplete data[20, 21].

In this study, we use SAEs to extract interpretable features from LLM embeddings for identifying ADRs. Our approach demonstrated strong classification performance across multiple health outcomes, suggesting that SAE-derived representations effectively retain critical information while simplifying model complexity. This process enhances interpretability and offers a promising route for improving pharmacovigilance applications.

## 2 Methods

### 2.1 Reference Data Source

We use a reference set of test cases from “Defining a Reference Set to Support Methodological Research in Drug Safety” by Ryan et al. [22]. A test case set in this context refers to pairs of drugs and symptoms, where the symptoms are specific adverse health outcomes. This reference set provides a benchmark across four key health outcomes: acute liver injury, acute kidney injury, acute myocardial infarction, and upper gastrointestinal bleeding.

The reference set comprises 399 test cases, including 165 positive controls (meaning **drug-adverse reaction pairs with established evidence of a causal relationship**) and 234 negative controls (meaning pairs with no evidence of a causal relationship). Positive controls are drug-symptom pairs supported by evidence from randomized clinical trials, observational studies, and case reports. Negative controls are selected based on the absence of evidence suggesting a causal relationship between the drug and the outcome.

### 2.2 Model Descriptions

#### Gemma-2-9b-it

Driven by the availability of pre-trained SAEs. we use the Gemma-2-9b-it[2] model developed by Google DeepMind. They trained the model on 8 trillion tokens, learning a diverse dataset that includes web documents, code, and scientific articles. The model’s architecture includes 42 layers with a model dimension of 3584 and a context length of 8192 tokens.

Gemma-2-9b-it specifically, is an instruction-tuned, meaning it has undergone posttraining fine-tuning to improve its performance on specific tasks, such as instruction following and safety. This tuning involves supervised fine-tuning and reinforcement learning from human feedback (RLHF), which helps align the model’s outputs with desired behaviors and reduces the likelihood of generating harmful content.

#### Gemma Scope

We use pre-trained SAEs from the Gemma Scope[23] project, specifically trained on Gemma2-9b-it’s residual stream after layers 9, 20, and 31, to transform the activations of drug names into interpretable features. The SAE expands the model from 3584 to 131,072 dimensions and uses a JumpReLU activation function, which enforce sparsity by zeroing out activations below a learned threshold, enhancing interpretability while maintaining reconstruction fidelity.

### 2.3 Classification Techniques for Probing

#### SAE-Driven Feature Extraction

Transforming hidden-state drug embeddings from the Gemma-2-9b-it model into interpretable features involves several key steps, as visually represented in Figure 1.

**Fig. 1.**
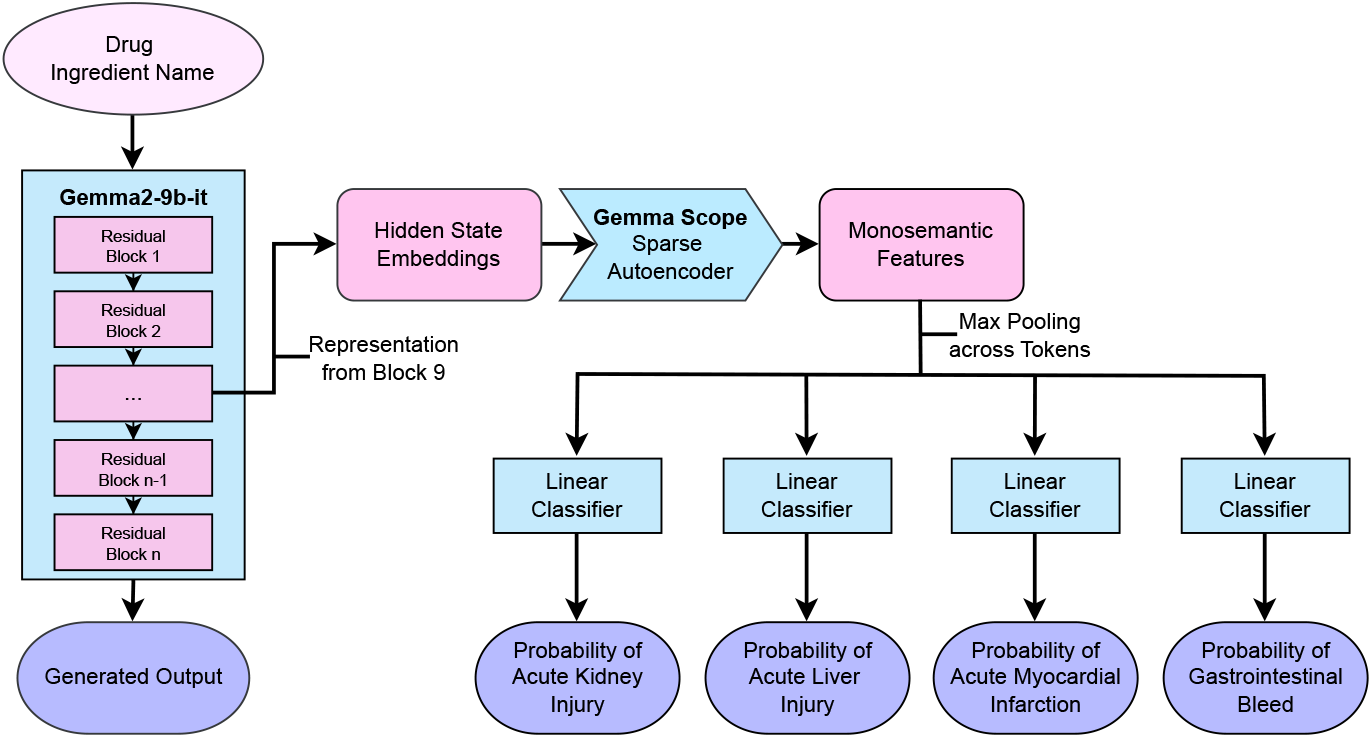
Flowchart for converting drug ingredient names into monosemantic features used to classify drug-ADR relationships.

We begin by extracting activations from a specific layer of the Gemma-2-9b-it model. These activations are processed using Gemma Scope to produce monosemantic features. The SAE transformation simplifies the complex embeddings, making them more interpretable while preserving critical information necessary for downstream analysis.

The transformed features are applied to our dataset of drug names associated with specific ADRs. Each drug name may be tokenized into *m* tokens, where each token *i* is represented by a feature vector 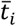. To manage the dimensionality and enhance interpretability, we condense these *m* token-level vectors via max pooling across tokens. Concretely, if each 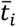 is *d*-dimensional, then the pooled vector 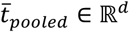 is computed by taking the maximum value in each dimension across all tokens:

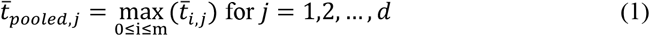

The pooled feature vector 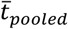 is then used as input to a logistic regression model to classify the likelihood of ADRs. Denote each dimension of 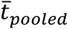 by *x*_*j*_. We predict the probability of an ADR occurring as follows:

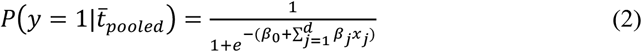

where β_0_ is the intercept, and β_j_ are the coefficients learned for each dimension *j*.

To encourage sparsity in the coefficients (which helps identify the most influential features), L1 regularization is applied. The penalized loss function is:

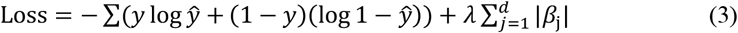

where λ is the L1 regularization parameter.

The model’s performance is evaluated using 10 repetitions of 5-fold cross-validation, and results are reported as area under the receiver operating characteristic (ROC) curve (AUC) scores across the selected health outcomes. To assess the statistical significance of the model’s performance, we conduct a permutation test[24], where the labels were shuffled and the AUC scores were recalculated to generate a distribution of scores under the null hypothesis. The p-values were computed by comparing the original AUC scores to this permuted distribution. Finally, to understand which pooled features are most indicative of ADR risk, we look at the difference between normalized features with nonzero L1-regularized coefficients.

#### Text Generation-Based Probing

To assess the generative capabilities of the Gemma-2-9b-it model in classifying ADRs, we worked with a straightforward prompting strategy. The model was prompted with the query:

*“<start_of_turn>user\nIs the drug {drug_name} known to cause {condition_name}? Answer using only ‘Yes’ or ‘No’*.*<end_of_turn>\n<start_of_turn>model\n*”

We then analyzed the probability assigned to the correct token (either “Yes” or “No”) as the next word. This probability was used to compute the ROC AUC.

#### Neural Networks on Unaltered Model Activations

To explore the predictive power of the unaltered model activations (3584 dense dimensions compared to the sparse 131,072 dimensions of our SAE activations), we implemented a shallow feed-forward neural network architecture. Specifically, after each residual block of the Gemma-2-9b-it model, we trained a multilayer perceptron (MLP) with a single hidden layer comprising 10 neurons on the residual stream. This setup was chosen to maintain simplicity while capturing essential patterns in the data. The model’s performance was evaluated using repeated stratified k-fold cross-validation, with 5 splits and 10 repetitions. Similarly to the other approaches, we looked at the AUC to evaluate performance.

## 3 Results

### 3.1 Classification Performance of Layer 9 Activations

The performance of the SAE-derived features from Layer 9 activations of the Gemma-2-9b-it model was evaluated for classifying ADRs. The SAE transformation of hidden-state embeddings of drug names resulted in monosemantic features that were subsequently used in a logistic regression model to classify ADRs.

For acute kidney injury, the area under the receiver operating characteristic curve (AUC) was 0.957, with a p-value of 0.020 from the permutation test. Similarly, for acute liver injury, the AUC was 0.902, with a p-value<0.001. The prediction of acute myocardial infarction yielded an AUC of 0.954, with a p-value<.001. Finally, the model achieved an AUC of 0.963 for gastrointestinal bleeds, with a p-value<.001. These results suggest that the SAE-derived features effectively capture the necessary information for accurate ADR classification.

Figure 2 illustrates the differences in expression values across the nonzero features identified by L1 regression with λ=.10.

**Fig. 2.**
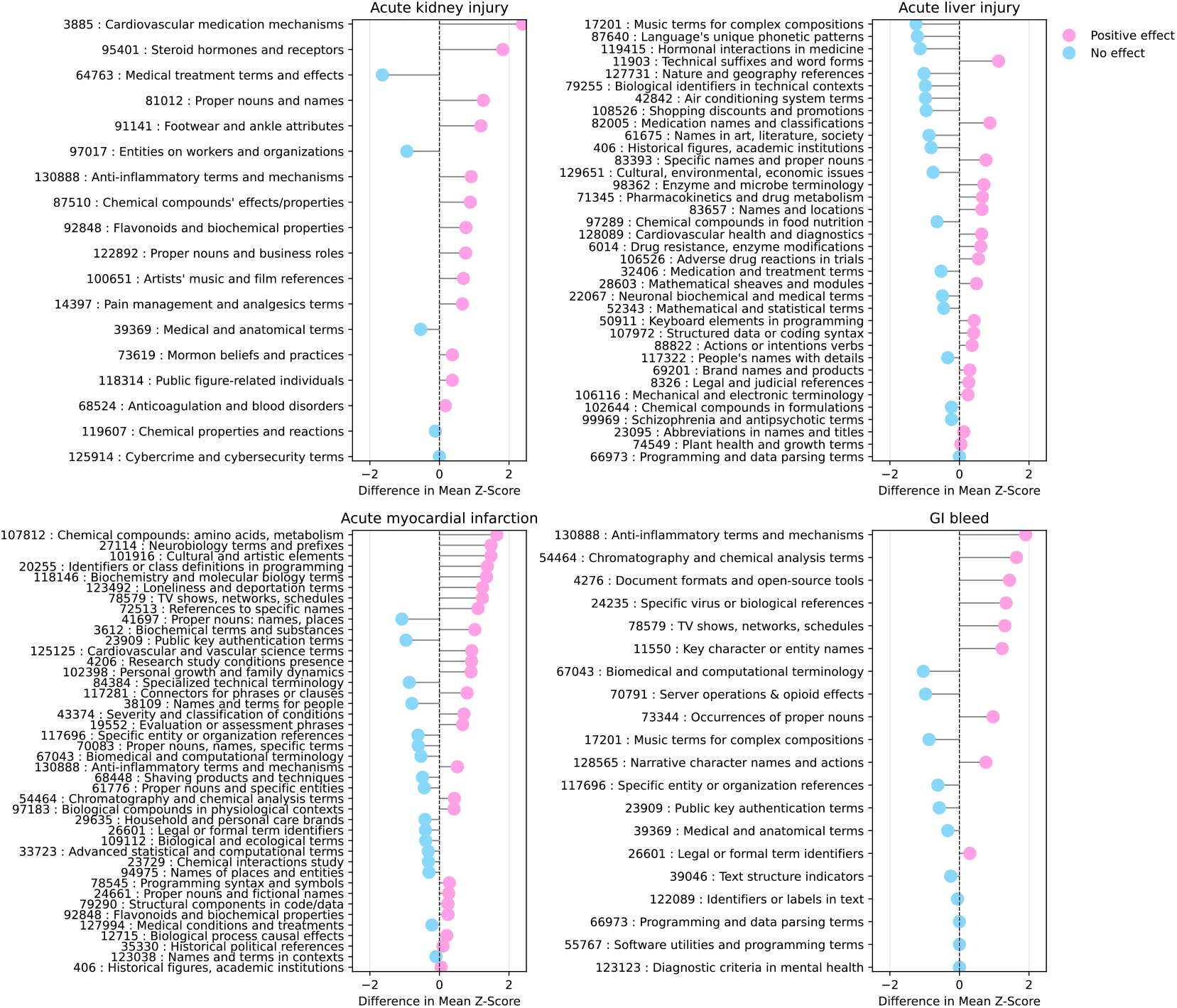
Lollipop plots showing the differences in mean Z-scores for nonzero sparse autoencoder features identified by L1 logistic regression across four conditions: acute kidney injury, acute liver injury, acute myocardial infarction, and gastrointestinal bleed. Feature descriptions are from https://www.neuronpedia.org/.

### 3.2 Comparison Across Layers and Approaches

We extend our analysis by evaluating SAEs trained on different layers of the Gemma-2-9b-it model, specifically layers 9, 20, and 31. Although pairwise t-tests among the three layers yield p-values of 0.016 for L9 vs L20, 0.884 for L9 vs L31, and 0.016 for L20 vs L31 (suggesting that L20 may differ statistically from the other two), the ranges of the AUC distributions in the boxplots overlap substantially. In practical terms, the performances across these layers are quite similar, with no clear advantage to choosing one layer over another based on these data alone.

We compare our classification SAE-derived features to a text-based generative approach, where the model was prompted to classify ADRs for a given drug directly. The generative method showed lower AUC scores compared to the SAE classifier on layer 9, with p-values indicating significant differences for each ADR: acute kidney injury (p = 0.001), acute liver injury (p = 0.789), acute myocardial infarction (p < 0.001), and gastrointestinal bleeds (p < 0.001). These results suggest that while the text generation-based approach captures some predictive information, the structured feature extraction via SAEs offers a more robust and interpretable solution.

Furthermore, we trained neural networks on the untransformed activations from each layer to explore their classification capabilities. These networks, evaluated across all layers, demonstrated varying performance. Figure 3 visualizes the comprehensive comparison of all methods, showing the AUC scores for SAEs, generative predictions, and neural networks across the evaluated layers.

**Fig. 3.**
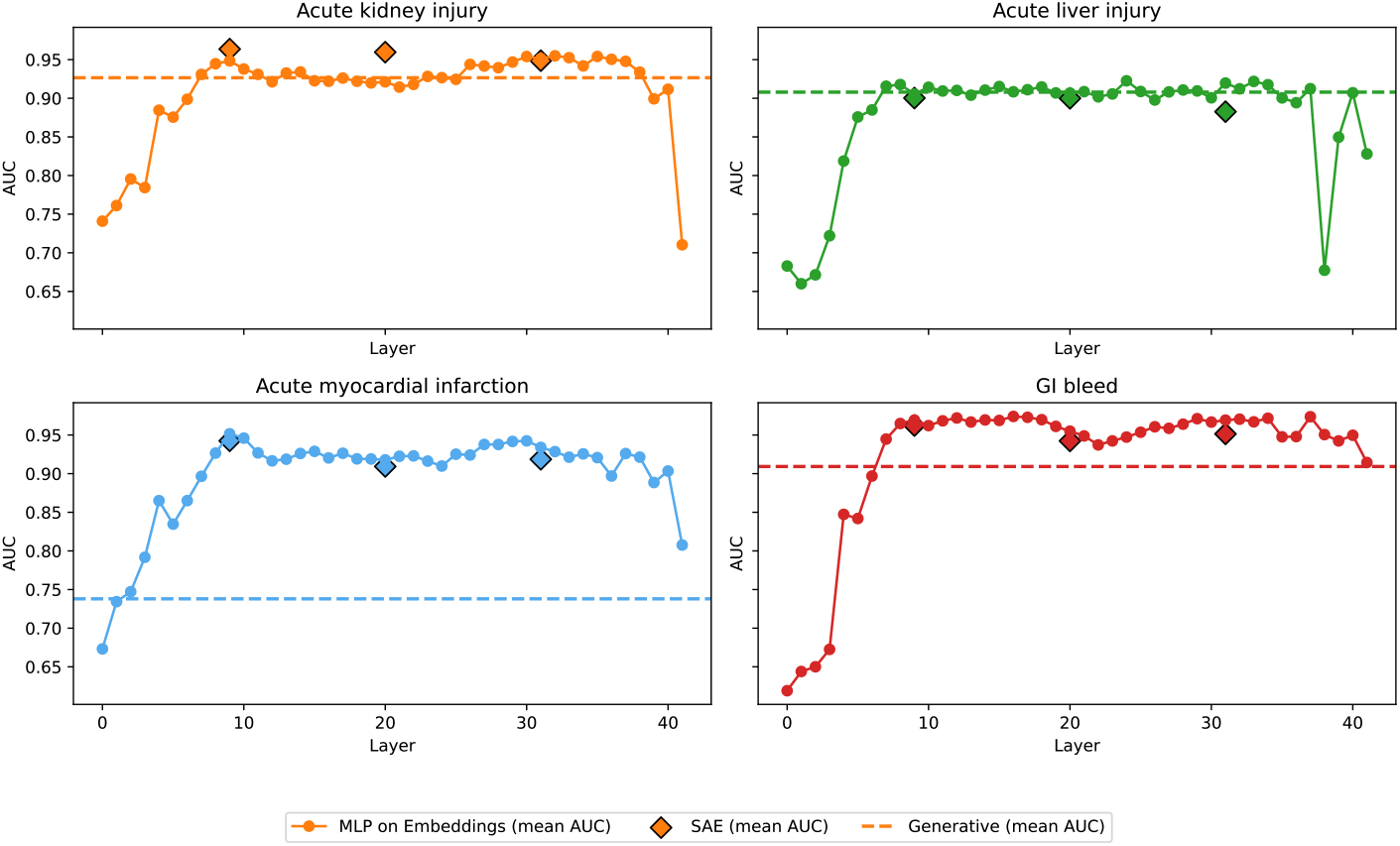
Comparison of mean AUC scores for ADR classification using SAE-derived features from layers 9, 20, and 31, generative predictions, and neural networks trained on raw activations across all layers.

## 4 Discussion

In this study, we explore how SAEs enhance the interpretability of the information within LLMs for ADR classification, revealing insights into the Gemma-2-9b-it model’s internal workings.

Examining the performance of early layers, particularly layers 0 to 9, we see that each layer demonstrates substantial gains in performance. These layers appear to capture foundational features that are crucial for identifying ADRs, suggesting that they hold generalizable patterns and representations. As we moved to later layers, we observed a plateau in performance, with the latest layers (40 to 42) showing sporadic results. This shift likely reflects the model’s focus on preparing for next-token predictions, a task that likely does not align well with the requirements for ADR classification.

Interestingly, a brief manual annotation revealed that only about half (46.6%) of the features identified by SAEs were somewhat science related. This partial monosemanticity suggests that while the features are not purely isolated to single concepts, they still activate in ways that are beneficial for ADR classification suggested by their retention through L1 regularization. For example, the feature 91141 (references to footwear or ankle attributes) appears in our layer 9 acute kidney injury model. We speculate that this may reflect broader associations with conditions that affect mobility or circulation, which are relevant to kidney health. Since features appear to split with an increase in SAE dimension[23], a larger SAE may better differentiate the relevant aspects of this feature.

The comparison between SAEs and the text-generation method revealed the advantages of structured feature extraction. While generative approaches rely heavily on prompt engineering, SAEs focus on internal representations. Prompt engineering can be thought of as optimizing the selection of features in the overall model structure. Learning from SAE activations does this automatically, without the trial and error of prompt engineering[25].

On this note, when comparing the SAE output to the unaltered activations, we found no significant differences in performance. This result is particularly encouraging, as it confirms that SAEs can simplify complex model outputs without losing critical information. This type of dictionary learning offers an interpretable approach to describing text input, which could simplify the circuitry of downstream machine learning tasks.

While our SAE-based approach shows promise in making the information within LLMs more interpretable for ADR classification, it’s important to acknowledge its limitations. The computational demands of training SAEs are intensive, potentially limiting their accessibility. Pretrained SAEs offer valuable insights into their parent LLMs, but training SAEs for additional models remains resource intensive[23]. Future research should focus on optimizing the training process or developing lightweight alternatives that maintain interpretability without the computational burden. Additionally, the partial monosemanticity observed in the extracted features suggests that there is room for improvement in achieving fully separable representations. Also worth noting, our generative approach was not heavily optimized through prompt engineering, which may have impacted its performance.

Looking ahead, future researchers could expand and refine our classification approach by exploring interactions and correlations between features. This would lead to a deeper understanding of how different features contribute to ADR classification and improve model accuracy. Additionally, using sparse crosscoders [26] to consider information across multiple layers could provide a more holistic view of the model’s internal representations, potentially uncovering new information on the complex dynamics within transformers.

Our findings indicate that the internal representations of LLMs, as interpreted through SAEs, provide a valuable synthesis of biomedical knowledge, as demonstrated in the context of ADR detection. By building upon these representations, we were able to validate known drug-ADR pairs, showcasing the potential for SAEs to interpret and utilize the information acquired during pretraining. This approach not only confirms existing knowledge but also opens the possibility of discovering new correlations that may not yet be recognized by experts. As we continue to explore these latent features, we aim to uncover novel insights that could enhance pharmacovigilance efforts and improve patient safety.

## Data Availability

All data produced are available online at https://pubmed.ncbi.nlm.nih.gov/24166222/

## Acknowledgments

NF is supported by NIH T32 HL116273.

