## Supplementary figures and images for "Probing Large Language Model Hidden States for Adverse Drug Reaction Knowledge"

### Supplemental Figure 1

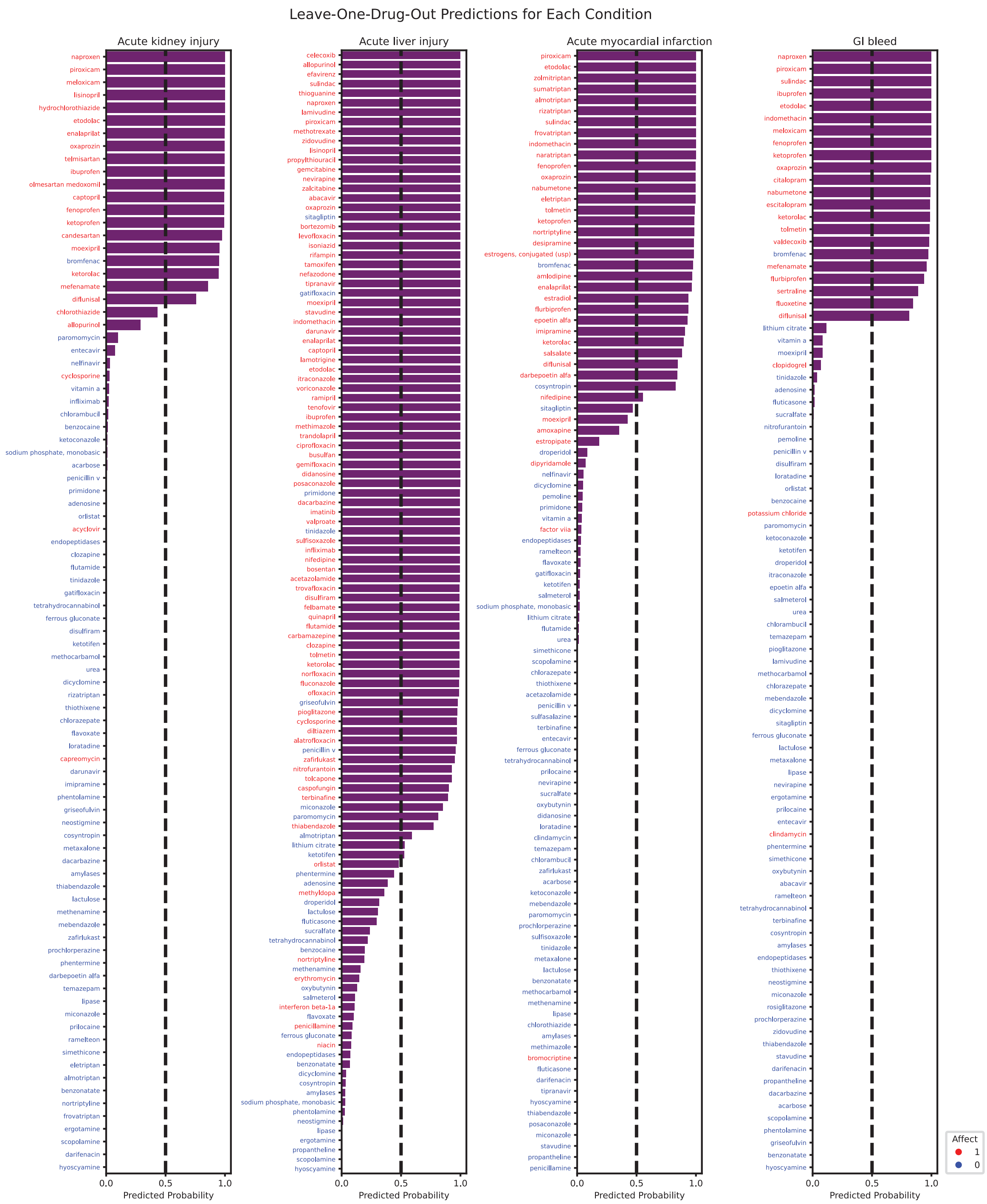
